# Cultural Stigma, Psychological Distress and Help-Seeking: Moderating role of Self-esteem and Self-stigma

**DOI:** 10.1101/2024.11.22.24317772

**Authors:** Toni Sawma, Souheil Hallit, Chloe Joy Younis, Jad Jaber, Myriam El Khoury-Malhame

**Affiliations:** Department of Psychology and Education, School of Arts and Sciences, Lebanese American University, Beirut, Lebanon; School of Medicine and Medical Sciences, Holy Spirit University of Kaslik, Lebanon; Research Department, Psychiatric Hospital of the Cross, Jal Eddib, Lebanon; Psychology Department, College of Humanities, Effat University, Saudi Arabia; Applied Science Research Center, Applied Science Private University, Amman, Jordan

**Author notes:** Corresponding Author Dr Myriam El Khoury-Malhame, Associate professor of psychology at the Department of Psychology and Education at the Lebanese American University.

**Keywords:** Mental disorders, stigma, Young adults, Lebanon, self-esteem

## Abstract

**Background/Objective:** Mental illness is a common and often stigmatized condition. Stigma around mental illness refers to the negative attitudes and beliefs society holds about individuals with mental health conditions, and can sometimes prevent those individuals from seeking adequate therapy. This study aims to explore the intricate relation between stigma, psychological distress, self-confidence and help-seeking behavior among young adults with diagnosed mental illnesses.

**Method:** A cross-sectional online survey was shared via digital platforms between February 2023 and August 2024. A total of 245 participants from various educational backgrounds and located in different Lebanese regions participated in the study and filled demographics data as well as assessments for stigma (Stigma Scale), self-esteem (Rosenberg Self-Esteem scale), help seeking (Attitudes Towards Seeking Professional Psychological Help -short form (ATSPPH-SF)), distress (Kessler Psychological Distress scale (K6)) and self-stigma (Self-Stigma Questionnaire (SSQ)).

**Results:** Results from our multivariable analysis revealed a significant association between high levels of stigma and low self-esteem scores, high levels of stigma and high psychological distress, as well as negative attitudes towards seeking professional help. We mostly found that fear of individuals with mental illnesses, prejudice and lack of enough knowledge and awareness were driving reasons for current stigma around mental health in Lebanon.

**Conclusion:** Stigma remains a pervasive condition leading to more mental health suffering, decreased self-esteem and lower tendencies to seek help. These inaccurate beliefs and stereotypes lead to fear, avoidance and overall discomfort, vis-à-vis people with mental distress perceived as unpredictable, impulsive and dangerous. It is important for public policy makers within collectivist culture to better address these misconceptions and promote help-seeking attitudes.

## BACKGROUND

In addition to the challenges of suffering from mental disorder(s), individuals often have to cope with the stigma that emerges from it as mental health stigma remains a widespread social problem impacting people’s well-being. According to the World Health Organization, stigma is associated with shame, embarrassment and rejection that causes an individual to be avoided or dismissed from others (WHO, 2019). When a phenomenon is stigmatized, a particular individual or group is labeled based on misconceptions and false stereotypes (Corrigan & Watson, 2004). The stigma associated with a particular condition, such as mental illness, can also result in social exclusion and discrimination (Link & Phelan, 2001). Furthermore, individuals who are subjected to stigma can experience loss of purpose and ambitions, loss of friendships and relationships as well as diminished quality of life and work opportunity threats (Lai et al., 2001). There is a significant body of research on stigma and its impacts, and scholars have investigated the different forms it can take. To begin with, the term social stigma refers to the public beliefs and impressions regarding a specific person or community (Watson et al., 2003). For example, perceiving an individual with schizophrenia as dangerous and crazy is a common illustration of social stigma. This might lead to internalized negative beliefs and perceptions that one who encounters stigma may develop about oneself are referred to as self-stigma (Link & Phelan, 2006). In fact, when individuals internalize societal perceptions, isolate themselves and feel embarrassed and ashamed of their mental illness, they are self-stigmatizing themselves. Finally, structural stigma, which emphasizes the institutional limitations, challenges and prejudice that people who suffer from mental health issues must overcome, especially in institutions and corporations (Pugh et al., 2015). One example of structural stigma might be restricting a person’s opportunities in the workplace because of their mental illness.

Among the most unfortunate outcomes of stigma around mental health is the threat of losing one’s self-esteem due to the ongoing instilled belief of being a disappointment and achieving nothing to be proud of (Link et al., 2001). According to numerous research (Wright et al., 2000; Link et al., 2001; Hayward et al., 2002), it has been documented that stigmatization has a negative impact on the self-esteem of those who are subjected to it since it leads to losing their sense of worth. This has been systematically reported in people who suffer from mental illness as stigma around their disorders correlates with lower self-esteem scores (Link and Phelan, 2006). In other words, individuals who have been labeled as “mentally ill” are led to believe that they are part of a social group not worthy of love and acceptance, incapable of conforming with others. Their anticipation of being discriminated against even intensifies underlying feelings of shame and guilt which in turn leads them to deny their sufferings, devaluate themselves, distort self-perceptions and worsen their self-esteem (Verhaeghe et al., 2008, Corrigan et al., 2006).

This decreased self-esteem due to stigma has been recently investigated in relation to inflating mental distress on one hand and hindering health seeking behaviors on the other (Quinn & Chaudoir, 2009, Gulliver et al. 2010). Global reports highlight extremely limited help-seeking behaviors in those struggling with mental distress with more than 75-80% of people with mental disorders not receiving therapy although approximately 1 in 4 people in Europe have a mental disorder (Clement et al., 2014). This is true even after massive collective traumas whereby only 10% of the people reported seeking mental support although made freely accessible by professionals after an urban disaster (El Khoury-Malhame et al., 2024). Pervasive stigma was reported as major indicator of unwillingness to access or continue mental health care services, in addition to financial considerations (Sirey et al., 2001, Hogan, 2003). This can leave those with mental diagnoses navigate challenges like relational complications, personal stress and anxiety (Livingston and Boyd, 2010, Vogel et al., 2006), as well as harmful coping mechanisms alone (Major & O’Brien, 2005). Social exclusion in turn decreases tendencies to seek help for stigmatized individuals with mental disorders (Gray, 2002). In fact, lack of social assistance is itself connected to high levels of sadness and anxiety (Cacioppo & Hawkley, 2003), and loneliness deteriorating altogether physical and mental conditions and prospective prognosis (Hawkley & Cacioppo, 2010). As such, individuals struggling with mental disorders additionally have to deal with extra layers of social ostracization and personal anxiety from being subjected to stigma around their mental health.

Minorities and youth are disproportionately deterred by stigma (Clement et al., 2014). It could be that for this vulnerable group, stigma extends beyond societal opinions and impacts one’s judgment of one’s own self (Villatoro et al., 2022). This key relationship between perceived stigma and personal stigma makes individuals with mental illnesses adopt negative attitudes towards themselves (Maeshima et al., 2022). Stigma causes negative self-perceptions and lower self-esteem levels, further burdening personal suffering inflating internalized negative stigma especially in young adults and adolescents (Ilic et al., 2012, Wood et al., 2017). Furthermore, this negative loop poses a barrier to getting professional psychological help or persevering with a prolonged treatment (Clement et al., 2015).

These widespread stigmatization around mental health seem even more problematic in the global South in general and Arab countries in particular, additionally hindering help-seeking (Fekih et al, 2022). Heightened stigma is partly accounted for by cultural and religious factors in this part of the world attributing mental health issues to supernatural causes (Ahad et al., 2023), in addition to poverty, lack of education and social shame or familial embarrassment (Jorm et al., 2012). Taken together, this sociocultural belief system can lead to increased discrimination and social exclusion of individuals with mental illnesses. In Lebanon, a small Levantine country, research on the influence of stigma on mental health and help-seeking behavior is scarce, although the country is going through major collective protracted crises. The protracted adversities have taken a toll on Lebanese youth mental health (El Khoury-Malhame et al., 2023), amidst unprecedented sociopolitical instability (Elkhoury-Malhame et al., 2024b). Lebanese with mental health issues reportedly feel stigmatized because of the cultural misconceptions around mental health, which has a detrimental effect on their self-esteem (Rayan and Fawaz, 2018). University students with mental illnesses further express emotions of shame and embarrassment, leading to their marginalization and social isolation (Wehbe, 2011). Stigma around mental health contributes to higher levels of distress among these young adults as they document significant levels of prejudice and greater feelings of despair and anxiety (Dimassi et al., 2015). Karam et al. (2019) highlights that people who suffer from mental disorders in Lebanon face major challenges when it comes to accessing mental health services and seeking psychological treatment because of the stigmatized association of seeking psychological help with signs of weakness, inadequacy and inability to deal with one’s own challenges. These constant social marginalization leads to internalized negative attitudes towards seeking assistance in young adults (Hassan, 2015), as they are fear cultural shaming of being called “crazy” (Al Laham et al., 2020).

The purpose of this research is thus to provide empirical data on the influence of stigma on self-esteem and help-seeking behaviors of diagnosed young adults in Lebanon. The objective is to better guide development of culturally and socially sensitive strategies to address the stigma associated with mental illness, and subsequently alleviate personal struggles, human suffering and socio-economic burden.

## METHODS

### Ethical Approval

In accordance with the Lebanese American University’s research protocol, the Institutional Review Board IRB approved this study (Tracking number LAU.SAS.MM2.8/Mar/2023). Participants electronically signed informed consents before enrolling.

### Study Design and Sample

The cross-sectional survey was conducted between March 2023 and August 2024. Participants responded to the survey link shared via social media platforms such as Instagram and WhatsApp. The sample consisted of 245 participants, with 147 women (60%). They ranged mostly between 20 and 29 years with 74.7% having a Bachelor’s degree. Participants were located in different Lebanese governorates and also professionally diagnosed with various types of mental illnesses.

### Procedure

Only participants who were above 18, living in Lebanon and diagnosed with at least one mental illness were eligible to participate. Participants were expected to complete the survey in 15 to 20 minutes. The number of participant responses that were excluded from the analysis was 14, and the cause of exclusion was due to incomplete survey responses.

### Measures

In addition to demographic data, participants were asked to fill the following scales:

#### The Stigma Scale

King et al. (2007) developed the 28-item stigma scale which is organized into three categories: discrimination, disclosure, and potential positive elements of mental illness. It is five-point Likert scale ranging from 1(strongly agree) to 5 (strongly disagree), with higher scores indicating greater stigma.

Some items include example of discriminatory conduct such as “I worry about telling people I receive psychological treatment”. This discrimination subscale measures the level of prejudice, exclusion, and discrimination experienced by individuals who suffer from mental illnesses. Moreover, the disclosure subscale measures the degree to which people with mental illnesses reveal their psychological struggles with statements like “People’s reactions to my mental health problems make me keep myself to myself”. The final subdivision of the scale measures the extent to which people with mental illnesses are seen as possessing positive traits like creativity and insight. This potential positive element subscale includes questions such as “My mental health problems have made me more accepting of other people”. The scale shows strong internal consistency and test-retest reliability with a Cronbach’s alpha of 0.87 for the discrimination subscale, 0.85 and 0.64 for the disclosure and potential positive elements subscales respectively. *The Rosenberg Self-Esteem Scale:* (Rosenberg, 1965). This scale consists of 10 items measuring self-esteem. It is a 4-point Linkert scale with response options varying from strongly agree to strongly disagree. Five items are positively worded and the other five are negatively worded. Higher scores suggest higher levels of self-esteem and self-worth. The scale has a Cronbach’s alpha of 0.84.

#### Attitudes Towards Seeking Professional Psychological Help Scale-short form (ATSPPH-SF)

(Fischer and Farina, 1995). It assesses social perceptions towards seeking psychological assistance. It includes 10 items on a 4-point Likert scale with statements such as “Personal and emotional troubles, like many things, tend to work out by themselves”. Responses range from 0 (disagree) to 3 (agree). Higher score indicates a more optimistic attitude and higher tendencies to ask for psychological aid. The scale had a Cronbach’s alpha of 0.84 among an Arab-speaking population (Rayan et al., 2020).

#### Kessler Psychological Distress Scale (Kessler et al., 2003)

A popular 6-item questionnaire used to measure psychological distress. It evaluates how frequently individuals have felt anxious, hopeless, or unworthy over the past 30 days. Each item is graded using a 5-point Likert scale, with the lowest score being 0 (none of the time) and 4 being the highest (all of the time), for a total final score that can range from 0 to 24. Higher scores mean higher levels of psychological distress. The K6’s internal consistency and reliability have been found to be high, with alpha coefficient being 0.86 (Ferro et al., 2019).

#### Self-Stigma Questionnaire SSQ

The 14-item questionnaire was developped to evaluate the degree to which people with mental illnesses internalize stereotypes and misconceptions, leading to self-stigmatization. Each item is evaluated on a 7-point Likert scale, with 1 (strongest agreement) and 7 (strongest disagreement). The scale measures how much individuals have absorbed and instilled unfavorable attitudes and beliefs about themselves and their psychological suffering, such as feelings of inferiority and shame because they have mental disorders. Three aspects of self-stigma are evaluated by the SSQ: internalized stigma, perceived discrimination and stereotypes as well as social functioning, with higher scores indicating lower levels of self-stigma. It has been confirmed to be reliable and effective across a range of populations and languages. high level of internal consistency, and Cronbach’s alpha coefficients ranging from 0.75 to 0.901 (Ochoa et al., 2015).

### Statistical Analysis

The SPSS software v.25 was used for the statistical analysis. The attitude help seeking score was considered normally distributed since the skewness (= .001) and kurtosis (= −.702) values varied between −1 and +1. The Student t was used to compare two means, the ANOVA test to compare three or more means and the Pearson test to correlate two continuous variables. The moderation analysis was conducted using PROCESS MACRO (an SPSS add-on) v3.4 model 1, taking the self-esteem, stigma and self-stigma scores as moderators between psychological distress and attitude help seeking. Results were adjusted over all variables that showed a p < .25 in the bivariate analysis. P < .05 was deemed statistically significant.

## RESULTS

### Sociodemographic and other characteristics of the sample

Two hundred forty-five young adults with diagnosed mental disorders participated in this study. Socio-demographics and descriptive statistics are represented in Table 1.

**Table 1.** Sociodemographic and other characteristics of the sample (N=245).

|  |  |
| --- | --- |
| **Variable** | **N (%)** |
| Age (years) |  |
| 18-19 | 16 (6.5%) |
| 20-29 | 220 (89.8%) |
| 30-39 | 7 (2.9%) |
| 40 and above | 2 (.8%) |
| Education |  |
| High school | 14 (5.7%) |
| Bachelor | 183 (75.0%) |
| Master | 46 (18.9%) |
| PhD | 2 (.8%) |

|  | **Mean ± SD** |
| --- | --- |
| Stigma (mean score) | 3.54 ± .81 |
| Self-esteem | 24.78 ± 2.37 |
| Attitude help seeking (mean score) | 2.26 ± .74 |
| Psychological distress (mean score) | 3.06 ± .93 |
| Self-stigma (mean score) | 2.93 ± 1.66 |

### Bivariate analysis of factors associated with attitude help seeking

The results showed that older participants and those with higher education compared to the other categories had significantly higher help-seeking attitude (Table 2). Higher stigma and psychological distress were significantly associated with lower help-seeking attitude, whereas having higher self-esteem and self-stigma were significantly associated with higher attitude help seeking (Table 3).

**Table 2.**
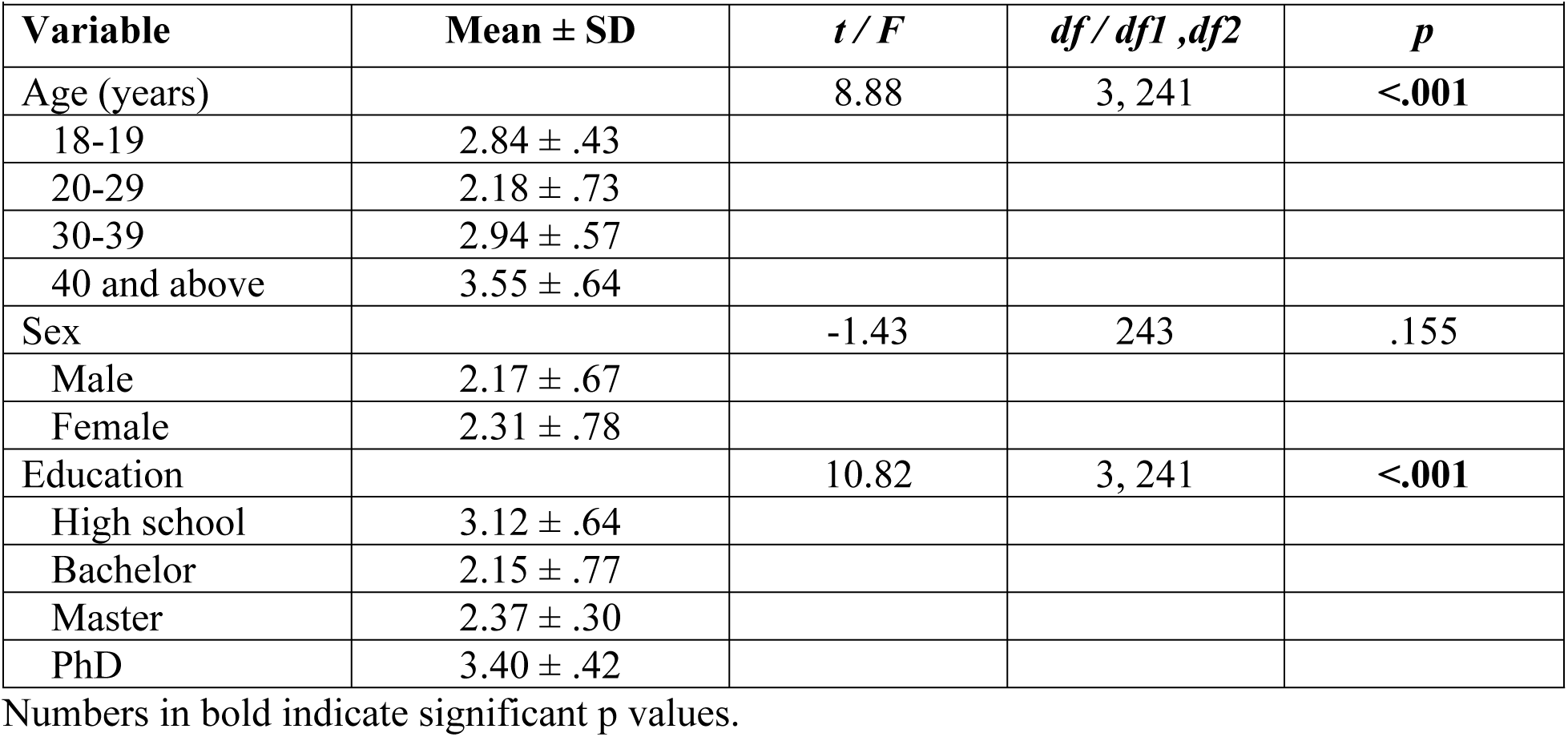
Bivariate analysis of factors associated with attitude help seeking.

|  |  |  |  |  |
| --- | --- | --- | --- | --- |
| **Variable** | **Mean ± SD** | ***t* / *F*** | ***df* / *df1* , *df2*** | ***p*** |
| Age (years) |  | 8.88 | 3, 241 | **<.001** |
| 18-19 | 2.84 ± .43 | | | |
| 20-29 | 2.18 ± .73 | | | |
| 30-39 | 2.94 ± .57 | | | |
| 40 and above | 3.55 ± .64 | | | |
| Sex |  | -1.43 | 243 | .155 |
| Male | 2.17 ± .67 | | | |
| Female | 2.31 ± .78 | | | |
| Education |  | 10.82 | 3, 241 | **<.001** |
| High school | 3.12 ± .64 | | | |
| Bachelor | 2.15 ± .77 | | | |
| Master | 2.37 ± .30 | | | |
| PhD | 3.40 ± .42 | | | |

Numbers in bold indicate significant p values.

**Table 3.**
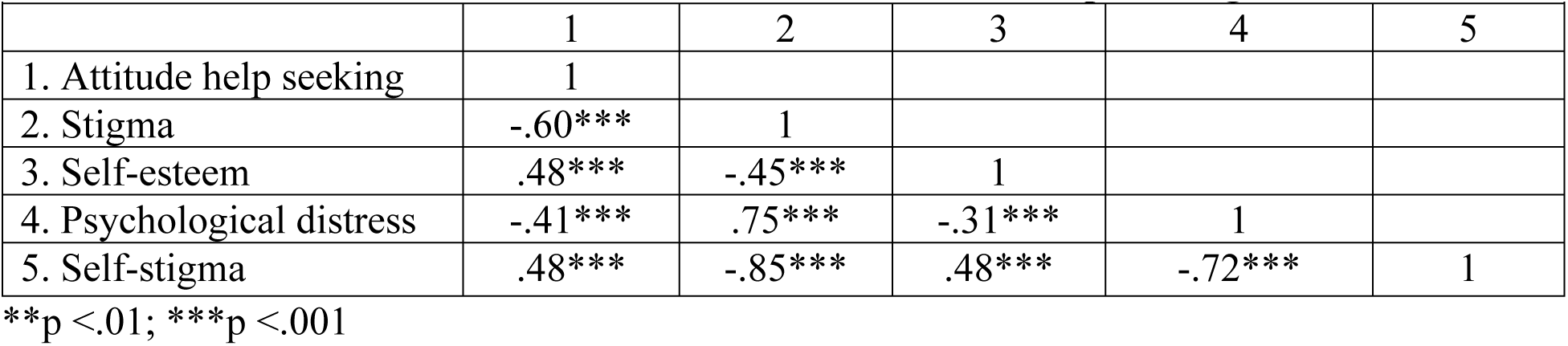
Correlations of continuous variables with attitude help seeking.

Numbers in bold indicate significant p values.
|  |  |  |  |  |  |
| --- | --- | --- | --- | --- | --- |
|  | 1 | 2 | 3 | 4 | 5 |
| 1. Attitude help seeking | 1 |  |  |  |  |
| 2. Stigma | -.60*** | 1 |  |  |  |
| 3. Self-esteem | .48*** | -.45*** | 1 |  |  |
| 4. Psychological distress | -.41*** | .75*** | -.31*** | 1 |  |
| 5. Self-stigma | .48*** | -.85*** | .48*** | -.72*** | 1 |

\*\*p <.01; \*\*\*p <.001

### Moderation analysis

The moderation models were adjusted over the following covariates: age, gender and education. The interaction self-esteem by psychological distress was significantly associated with help-seeking attitude (Beta= .08; t= 4.07; p < .001; 95% CI .04; .12) (Table 4, Model 1).

**Table 4.** Moderation analyses taking attitude help seeking as the dependent variable.

|  |  |  |  |  |
| --- | --- | --- | --- | --- |
|  | **Beta** | ***t*** | ***p*** | **95% CI** |
| Model 1: Self-esteem as the moderator |  |  |  |  |
| Psychological distress | -2.19 | -4.49 | **<.001** | -3.15; -1.23 |
| Self-esteem | -.11 | -1.94 | .054 | -.23; .002 |
| Interaction psychological distress by self-esteem | .08 | 4.07 | **<.001** | .04; .12 |
| Model 2: Self-stigma as the moderator |  |  |  |  |
| Psychological distress | -.37 | -3.34 | **.001** | -.58; -.15 |
| Self-stigma | -.03 | -.44 | .658 | -.18; .11 |
| Interaction psychological distress by self-stigma | .07 | 2.68 | **.008** | .02; .12 |

Numbers in bold indicate significant $p$ values.

At low (Beta = −.44; *p* <.001) and moderate (Beta = −.25; *p* <.001) levels of self-esteem, higher psychological distress was significantly associated with lower help-seeking attitude (Table 5, Model 1).

**Table 5.** Conditional effects of the focal predictor (psychological distress) at values of the moderator.

Numbers in bold indicate significant $p$ values.
|  |  |  |  |  |
| --- | --- | --- | --- | --- |
|  | **Beta** | ***t*** | ***p*** | **95% CI** |
| Model 1: Self-esteem as the moderator |  |  |  |  |
| Low (= 22.41) | -.44 | -6.19 | **<.001** | -.57; -.30 |
| Moderate (= 24.78) | -.25 | -5.52 | **<.001** | -.34; -.16 |
| High (= 27.15) | -.06 | -1.10 | .273 | -.18; .05 |

| Model 2: Self-stigma as the moderator |  |  |  |  |
| --- | --- | --- | --- | --- |
| Low (= 1.27) | -.28 | -3.28 | **.001** | -.45; -.11 |
| Moderate (= 2.93) | -.17 | -2.55 | **.011** | -.29; -.04 |
| High (= 4.59) | -.05 | -.75 | .455 | -.19; .08 |

Numbers in bold indicate significant $p$ values.

Also, the interaction self-stigma by psychological distress was significantly associated with help-seeking attitude (Beta= .07; t= 2.68; p = .008; 95% CI .02; .12) (Table 4, Model 2).

At low (Beta = −.28; *p* = .001) and moderate (Beta = −.17; *p* = .011) levels of self-stigma, higher psychological distress was significantly associated with lower help-seeking attitude (Table 5, Model 2).

## DISCUSSION

The impact of stigma on mental health is a major public health issue with cultural sensitivities and is best understood when including individuals with diagnosable disorders. In our study, stigma around mental illness as shown to deter young adults’ mental distress and further defer them from seeking recovery in Lebanon. The relationship between stigma, psychological distress and help-seeking intentions is further complexified and influenced by both self-esteem and self-stigma, that are shown to play crucial mediating roles respectively.

### Help-Seeking in relation to Stigma and Psychological Distress

Our findings first and foremost support accumulating evidence that stigma very significantly correlates with increased psychological distress, self-stigma and deteriorated self-esteem while impairing help-seeking. This is in line with recent data conducted in the same context among Lebanese individuals with mental illnesses, whereby participants reported lower levels of self-esteem and quality of life, as well as higher levels of social isolation when subjected to stigma (El Hayek et al., 2021). Our results further echo the extent to which stigma inflates feelings of humiliation and embarrassment, leading to negative self-perceptions. Heightened levels of stigmatization can increase psychological distress symptoms of anxiety, depression (Burgess et al., 2019) all while increasing self-stigma and loss of self-worth, leading to hopelessness and helplessness (Corrigan and Watson, 2002). Individuals become even less likely to seek professional help (Vogel et al., 2006), even when mental health services are accessible and affordable (Sirey et al., 2001). Accumulating evidence support the negative impact of self-stigma as a barrier to seeking mental health services in young adults already suffering from mental illness (Schomerus & Angermeyer, 2008, Jennings et al. 2015). They indeed seem to synchronize their disbelief with their own competences as they internalize negative misbeliefs and misconceptions with the societal pervasive stigma associated with their mental illness. This reinforces marginalization and social isolation and perpetuates prevalent stigma and discrimination (Link & Phelan, 2001). It also solidifies barriers to seeking mental health recovery in occidental (Yanos et al., 2010) as well as oriental societies (Obeid et al., 2019) as people suffering from mental illnesses fear of prejudice and discrimination of seeking treatment. In the Lebanese context, self-stigma is still a prevalent challenge and mental health is still frowned upon (Dalky, 2012).

Individuals’ confidence in getting the proper care might be additionally hindered by cultural assumptions that psychological disorders are personal failings rather than conditions that need to be recognized and treated (Karam et al., 2019, Krstanoska-Blazeska & Slewa-Younan 2021). Education and older age seem to play protective factors as we found that older adults in the sample and those with highest degrees had considerably more positive attitudes towards help-seeking behavior. Charara et al. (2017) had previously demonstrated that lack of education and awareness contributed to limited understanding of mental health and to subsequent stigma. Highlighting and targeting such social and personal stigmas could thus be considered as a public health priority since studies suggest that more favorable attitudes on seeking mental health professional help are linked to improved levels of self-esteem and restituted social functioning (Vogel et al., 2006).

### Help-Seeking in relation to Self-Esteem

Our results suggest that individuals with low to moderate self-esteem, who are already vulnerable to psychological distress, are less likely to seek help for their mental health concerns. This negative correlation between self-esteem and help-seeking attitudes highlights the detrimental role of stigma in exacerbating mental health difficulties. Individuals with low self-esteem may internalize societal stigma, leading to feelings of shame, guilt, and inadequacy, which can further hinder their willingness to seek professional help. In fact, individuals with high levels of self-esteem and self-worth generally describe help-seeking as empowering and beneficial rather than a sign of weakness and vulnerability (Lannin et al., 2015). University students with higher self-esteem seem to be queen on promptly recognizing and addressing issues related to mental health for better outcomes and faster recovery (Yih, 2020). These results underscore the importance of addressing stigma and promoting self-esteem to encourage help-seeking behaviors among young adults with mental disorders.

### Help-Seeking in relation to Self-Stigma

This study also illustrates the role self-stigma in moderating the relationship between psychological distress and help-seeking intentions. Self-stigma refers to students’ internalization of cultural mental health stigmas and feel like a disgrace and disappointment to their families. Adopt negative perceptions and stereotypes, makes feelings of worthlessness, shame and guilt emerge, reinforcing unfavorable perceptions toward mental illnesses (Cheng, 2019). The interaction effect between self-stigma and psychological distress further underscores the complexity of help-seeking behaviors. Specifically, individuals with low to moderate levels of self-stigma who also experience high psychological distress are less likely to seek help. This finding suggests that self-stigma can amplify the negative impact of psychological distress on help-seeking attitudes. As self-stigma increases, individuals may become more reluctant to acknowledge their mental health problems, fear negative judgment, and avoid seeking necessary support. Within this framework, self-esteem may serve as a protective factor, mitigating the negative effects of both self-stigma and psychological distress on help-seeking intentions. Individuals with higher self-esteem may be more resilient to the negative impacts of stigma and distress, making them more likely to seek help. These positive attitudes seem better supported with the positive assistance of healthcare providers (Abi Hana et al., 2022).

### Limitations and Future Research

Increasing the sample size could benefit in more adequate generalization of the findings. Qualitative reports that specifically target the Lebanese cultural context could provide additional support to some sensitive aspect of stigma and would better encompass the range of diversity and experiences within different Lebanese communities related to gender, religious or urban versus rural settings. Conducting a longitudinal design and including therapy outcome could also track potential fluctuations in stigma, self-stigma, self-esteem, psychological distress and help-seeking tendencies across several mental conditions. Lastly, it could be useful to track the psychiatric diagnoses of participants to overcome some social desirability biases when self-reporting mental disorders.

## Conclusion

In conclusion, this study sheds light on the significance of mental health stigma on psychological distress and help-seeking. Findings highlight the pivotal role of self-esteem and self-stigma in moderating the above intricate relations in a collectivist crisis enriched context. This builds ontop of recommendations from the Lancet Commission on ending stigma and discrimination in mental health (2022). If policy makers capitalize on positive stigma awareness and incentivize engagement in proactive behaviors towards destigmatizing and treating mental illness this could increase help-seeking attitudes and behaviors, with favorable outcomes on mental and physical health.

## Declarations

### Human Ethics approval and Consent to participate declaration

In accordance with the Declaration of Helsinki, all participants approved and signed the informed consent as per the IRB ethics committee approval and tracking number LAU.SAS.MM2.8/Mar/2023

### Availability of data and materials

The datasets used and/or analyzes during the current study are available from the corresponding author on reasonable request.

### Competing interests

The authors declare that they have no competing interests

## Funding

Not Applicable

## Authors’ contributions

M.EK and T.S designed the study and wrote the manuscript. C. Y and J.J collected data and drafted part of the main manuscript and S.H prepared the analyses and corresponding tables. All authors reviewed the manuscript.

## Data Availability

Data are not currently shared publicly. They will be made available for researchers upon reasonable request

## Acknowledgements

The authors wish to acknowledge all those who made the project possible.

## Notes

### Competing Interest Statement

The authors have declared no competing interest.

### Funding Statement

The author(s) received no specific funding for this work.

### Author Declarations

In accordance with the Lebanese American University's research protocol, the Institutional Review Board IRB approved this study (Tracking number LAU.SAS.MM2.8/Mar/2023).

